# Performance of Off-the-Shelf Machine Learning Architectures and Biases in Detection of Low Left Ventricular Ejection Fraction

**DOI:** 10.1101/2023.06.10.23291237

**Authors:** Jake A. Bergquist, Brian Zenger, James Brundage, Rob S. MacLeod, T. Jared Bunch, Rashmee Shah, Xiangyang Ye, Ann Lyons, Ravi Ranjan, Tolga Tasdizen, Benjamin A. Steinberg

**Affiliations:** Scientific Computing and Imaging Institute, University of Utah, SLC, UT, USA; Nora Eccles Treadwell CVRTI, University of Utah, SLC, UT, USA; Department of Biomedical Engineering, University of Utah, SLC, UT, USA; School of Medicine, University of Utah, SLC, UT, USA; Data Science Services, University of Utah, SLC, UT, USA; Department of Electrical & Computer Engineering, University of Utah, SLC, UT, USA

## Abstract

Artificial intelligence - machine learning (AI-ML) is a computational technique that has been demonstrated to be able to extract meaningful clinical information from diagnostic data that are not available using either human interpretation or more simple analysis methods. Recent developments have shown that AI-ML approaches applied to ECGs can accurately predict different patient characteristics and pathologies not detectable by expert physician readers. There is an extensive body of literature surrounding the use of AI-ML in other fields, which has given rise to an array of predefined open-source AI-ML architectures which can be translated to new problems in an “off-the-shelf” manner. Applying “off-the-shelf” AI-ML architectures to ECG-based datasets opens the door for rapid development and identification of previously unknown disease biomarkers. Despite the excellent opportunity, the ideal open-source AI-ML architecture for ECG related problems is not known. Furthermore, there has been limited investigation on how and when these AI-ML approaches fail and possible bias or disparities associated with particular network architectures. In this study, we aimed to: (1) determine if open-source, “off-the-shelf” AI-ML architectures could be trained to classify low LVEF from ECGs, (2) assess the accuracy of different AI-ML architectures compared to each other, and (3) to identify which, if any, patient characteristics are associated with poor AI-ML performance.

## 2 Introduction

Artificial intelligence - machine learning (AI-ML) is a computational technique that has been demonstrated to be able to extract meaningful clinical information from diagnostic data that are not available using either human interpretation or more simple analysis methods.[1]–[5] AI-ML has demonstrated remarkable successes across many clinical domains, including the 12-lead electrocardiogram (ECG) [4], [6], [7]. Recent developments have shown that AI-ML approaches applied to ECGs can accurately predict different patient characteristics and pathologies not detectable by expert physician readers, including age, sex, and low left ventricular ejection fraction (low LVEF) [4], [6]–[8]. Some AI-ML tools pending FDA authorization are being implemented in medical systems as diagnostic tests that can run on collected ECGs and provide additional diagnostic information [4], [6], [7].

AI-ML, while relatively new in the healthcare space, has been around for several decades. Many robust AI-ML tools have been designed and applied across various problems, including image analysis, text prediction, and “chatbots” such as chat-GPT[9]. The progression of these tools has gone through revolutionary development with a thriving community creating “open-source” or freely available, predesigned AI-ML architectures that can be easily trained on similar classification problems, “off-the-shelf” [10]. However, these architectures have not been robustly applied to ECG analysis. Many of the AI-ML techniques currently used in ECG analysis use custom implementations, which limits the trust and portability of these tools applied to other ECG datasets from different patient populations [4], [11]–[14]. Custom and proprietary AI-ML architectures also inhibit the development and collaborative improvement of these approaches applied to relevant clinical problems. Applying “off-the-shelf” AI-ML architectures to ECG-based datasets opens the door for rapid development and identification of previously unknown disease biomarkers.

Despite the excellent opportunity, the ideal open-source AI-ML architecture for ECG related problems is not known. Furthermore, there has been limited investigation on how and when these AI-ML approaches fail and possible bias or disparities associated with particular network architectures. In this study, we aimed to: (1) determine if open-source, “off-the-shelf” AI-ML architectures could be trained to classify low LVEF from ECGs, (2) assess the accuracy of different AI-ML architectures compared to each other, and (3) to identify which, if any, patient characteristics are associated with poor AI-ML performance.

## 3 Methods

### 3.1 ECG Dataset

As part of data associated with routine clinical care, ECGs with and LVEF measurement data were extracted from the University of Utah Electronic Data Warehouse from 2012-2021, resulting in 24,868 unique patient-ECG pairs. For this study, patients with an ECG recording within 30 days (average of 4.4 days ± 7.3 days) of an LVEF measurement via echocardiogram were selected. LVEF was calculated using echocardiography measurements verified by board-certified cardiologists. Low LVEF was defined as below 40%. The ECG recordings, performed on a GE-Marquette ECG Machine (Marquette, WI), included Leads I, II, and v1–6. Leads III, aVF, aVR and aVL can each be derived from the remaining leads, and thus are not use in such analyses. As is standard, recordings were 10 seconds long and sampled at 500Hz, resulting in an 8 × 5000 point array for each ECG. Patients were split into a 90% training (22,382 patients) and 10% (2,486 patients) testing set. The same training and testing set was used for all analyses. A summary of the patient characteristics for the training and testing set is presented in Table 2.

**Table 1:**
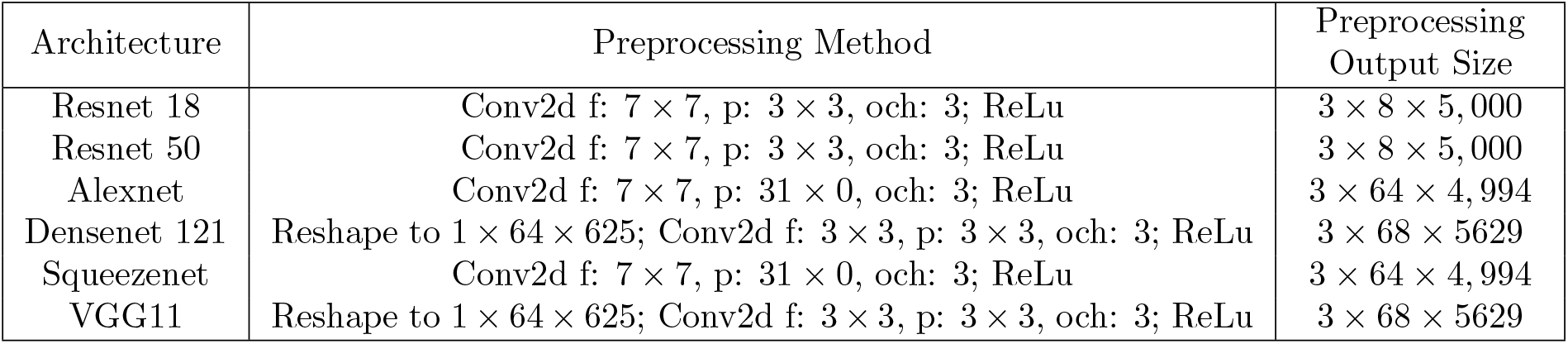
Preprocessing of ECG signals for each off-the-shelf network. Reprocessing consisted of a combination of reshaping (if necessary) of the 1×8×5, 000 (channels by electrodes by time) ECG input, a 2d convolutional layer (filter size f:*a × b*, padding p:*c × d*, and three output channels (och)) followed by a ReLu layer.

**Table 2:** Patient demographics in the training and testing sets. Statistical significance indicates a difference between training and testing patients.

| Variable | Test N=2486 | Train N=22382 | P value | x Missing |
| --- | --- | --- | --- | --- |
| **Female** | 1073 (45.31%) | 9694 (45.43%) | 0.91 | 1162 |
| **Race:** White or Caucasian | 1948 (82.65%) | 17569 (82.59%) | 0.84 | 1238 |
| Black or African American | 54 (2.29%) | 468 (2.2%) | — | — |
| Asian | 44 (1.87%) | 376 (1.77%) | — | — |
| American Indian and Alaska Native | 45 (1.91%) | 356 (1.67%) | — | — |
| Other Pacific Islander | 27 (1.15%) | 298 (1.4%) | — | — |
| Unknown | 37 (1.57%) | 281 (1.32%) | — | — |
| Other | 184 (7.81%) | 1751 (8.23%) | — | — |
| Choose not to disclose | 18 (0.76%) | 174 (0.82%) | — | — |
| **Hypertension** | 583 (24.66%) | 5139 (24.14%) | 0.57 | 1216 |
| **Diabetes** | 627 (26.52%) | 5709 (26.82%) | 0.76 | 1216 |
| **Obstructive Sleep Apnea** | 323 (13.66%) | 3071 (14.43%) | 0.32 | 1216 |
| **Cancer** | 384 (16.24%) | 3775 (17.73%) | 0.07 | 1216 |
| **Chronic Kidney Disease** | 389 (16.46%) | 3331 (15.65%) | 0.31 | 1216 |
| **Liver Disease** | 271 (11.46%) | 2488 (11.69%) | 0.75 | 1216 |
| **Chronic Obstructive Pulmonary Disease** | 595 (25.17%) | 5341 (25.09%) | 0.93 | 1216 |
| **Dementia** | 62 (2.62%) | 558 (2.62%) | 1.00 | 1216 |
| **Depression** | 659 (27.88%) | 5863 (27.54%) | 0.73 | 1216 |
| **Peripheral Artery Disease** | 305 (12.9%) | 2584 (12.14%) | 0.28 | 1216 |
| **Cardiac Valve Disease** | 394 (16.67%) | 3627 (17.04%) | 0.65 | 1216 |

### 3.2 Patient Characteristics

Based on our previously described methodology, clinical data were derived from the healthcare system’s enterprise data warehouse and include all administrative billing encounters with diagnosis codes (inpatient, outpatient, procedural), medication orders, and laboratory results [15], [16]. Clinical comorbidities were measured using previously validated algorithms in administrative data analyses of cardiovascular disease and included all healthcare system encounters up to and including the index visit. Comorbidity rates were calculated based on ICD codes as part of clinical billing encounters, as previously described [15], [17]. The index visit was defined as the date of echocardiogram acquisition.

### 3.3 Machine Learning Architectures

Open-source ML architectures from the Pytorch python-based machine learning package were adapted with ECG inputs. Specifically, we implemented untrained versions of Resnet 18, Resnet 50, Allexnet, Densenet 121, Squeezenet 1 0, and VGG 11 (https://pytorch.org/vision/stable/models.html) [10]. Each network architecture was developed for use with images and by default required a 3 *× m × n* input tensor (channels × height × width) and produced a 1,000 feature vector output. To minimally adapt these architectures to ECG signals, which consist of only one channel, eight signals, and five thousand time instances (1 × 8 × 5, 000), we preprocessed the input ECGs by adding a 2D convolutional layer to the beginning of each network with three output channels. To process the output 1,000 features we then appended a final fully connected layer with a single feature output followed by a rectified linear unit (ReLU) layer as the network output. Because of the architectures of the open-source implementations, in some cases it was necessary to either zero pad ECGs or restructure the input ECG to prevent a collapse in the lead dimension. The overall network design for adapting the open-source architectures for use with ECG data is depicted in Figure 1. Table 1 details the specific preprocessing steps used for each AI-ML architecture. No augmentations or transformations were performed on the input ECGs other than those listed in Table 1.

**Figure 1:**
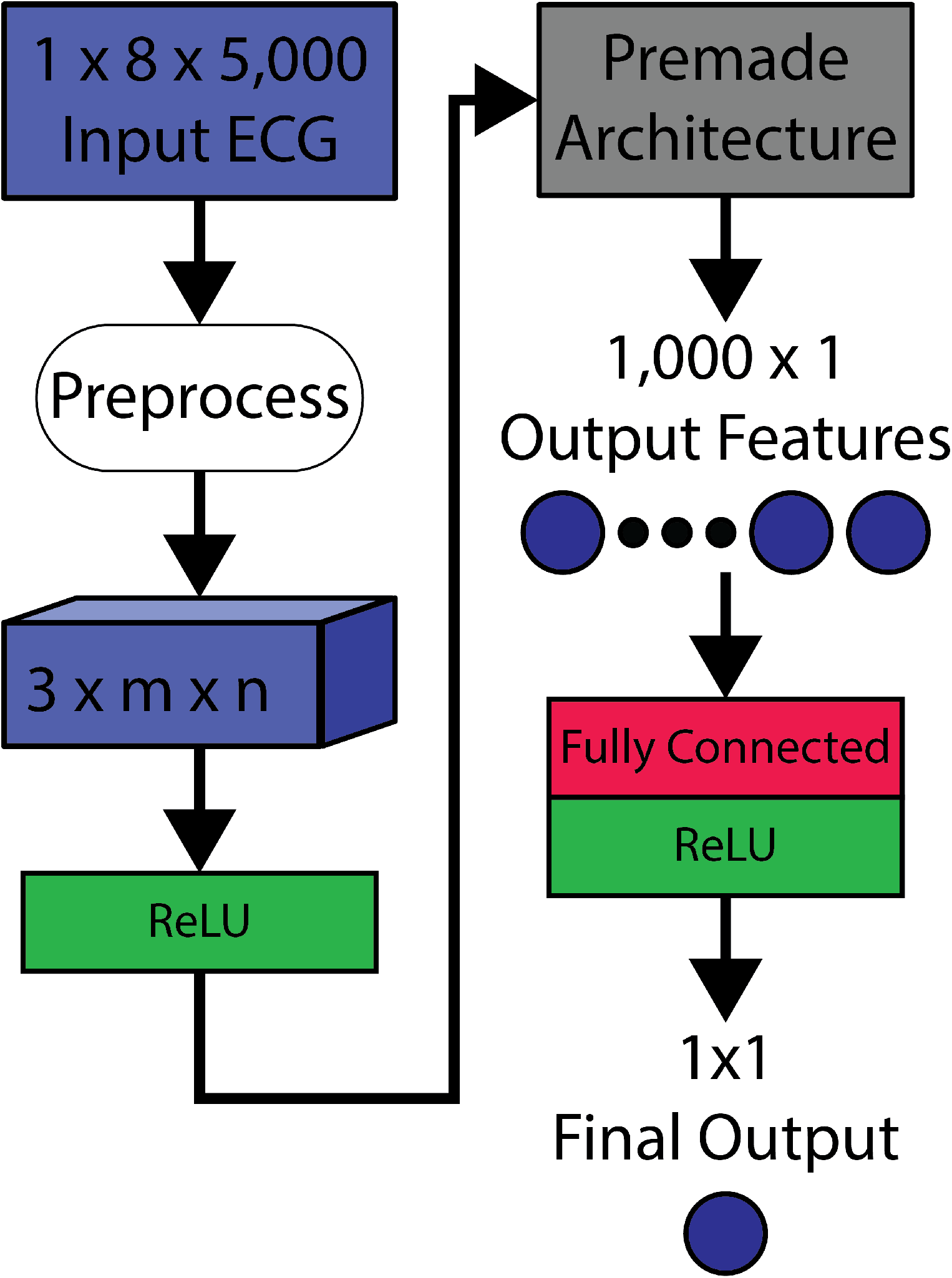
General infrastructure to adapt off-the-shelf ML architectures to operate on ECG data. Input ECGs are considered as a 1 channel, 8 lead, 5,000 time instant tensor (1 × 8 × 5, 000 input). The input ECGs are then preprocessed through a combination of reshaping (if needed), padding (if needed), and an initial convolutional layer to produce a 3 channel preprocessed tensor (3 *× m × n*). The preprossessing steps for each premade architecture are detailed in Table 1. The data is then passed through a rectified linear unit (ReLU) before entering the premade architecture. The output of the premade architecture is a 1 × 1, 000 output feature vector. These features are passed through a fully connected layer and another ReLU to produce a single output value.

Each network was trained using an Adam optimizer and binary cross-entropy loss between the network output and target LVEF classification [5]. AUC for the test dataset was monitored throughout training, and the network weights that produced the highest test AUC were saved to prevent over-fitting to the training set. The training was continued for 50 iterations, and the time to complete all iterations was recorded. Weights and biases were initialized randomly for each network. For each AI-ML architecture, five separate instances were trained to account for differences caused by random initialization of the network weights and biases.

### 3.4 AI-ML Performance Analysis

Each trained network (five instances per network architecture for a total of 30 networks) was evaluated on the testing dataset according to a range of standard metrics, including area under the receiver operator curve (AUC), F1 score, sensitivity, and specificity. The output of each architecture is a continuous variable between 0 and 1 that must be thresholded to produce a binary classification of low LVEF. To identify a robust threshold for each network, we selected a threshold that produced the highest F1 score in each network architecture. This threshold was used for the calculation of specificity and sensitivity for each network. Next, using the best performing instance (highest AUC) per network architecture, we grouped patients into incorrect prediction (false negative or false positive) or correct prediction (true negative or true positive) groups for each architecture. These groups were then used in subsequent demographic and comorbidity analysis.

### 3.5 Clinical Comorbidity Analysis

We computed descriptive statistics and summarized the distribution of patient demographic characteristics and medical conditions for numeric and categorical variables. Univariate comparisons across all patient characteristics between correct vs. incorrect LVEF classifications were performed for each network architecture. For these comparisons, the best-performing instance of each AI-ML architecture was used to group correctly vs. incorrectly classified patients.

Data processing was performed using R (Version 3.6.3), and RStudio (Version 1.2.5033), with appropriate packages. Statistical analysis was performed using R (Version 4.1.0) and RStudio (Version 1.0.153)[18]–[20].

Analysis of the data collected as part of routine clinical care, and subsequent reporting of anonymized, aggregate data, was approved by the University of Utah Institutional Review Board. The Institutional Review Board waived consent because the study is a retrospective analysis with minimal patient risk. The research reported adheres to the Helsinki Declaration guidelines on human research.

### 3.6 Computational Implementation

All ML architectures were implemented in Pytorch, an open-source AI/ML library.[10] Models were trained and evaluated on a system consisting of one NVIDIA TITAN RTX graphic card (24 GB video ram, CUDA 11.4), two Intel(R) Xeon(R) Silver 4114 CPUs at 2.20GHz (20 cores total), 256 GB DDR4 memory, Open Suse Leap version 15.0.

## 4 Results

### 4.1 Network Performance

All networks were trained using 22,382 combined ECG-LVEF pairs. Network performance was tested using 2,486 ECG-LVEF pairs. Baseline patient characteristics and comorbidities can be found in table 2.

The performance of each AI-ML architecture is shown in figure 2. Resnet 18 was, on average, the highest-performing architecture with a mean AUC of 0.917 ± 0.001. VGG 11 was the worst-performing architecture with an AUC of 0.902 ± 0.004. F-score was also used to assess network accuracy. The maximum F1 score was computed for each network by testing a range of network output thresholds. Resnet 18 showed the highest performance with an average max F1 score of 0.586 ± 0.010, and VGG 11 had the lowest with 0.520 ± 0.010. The threshold corresponding to the maximum F1 score was used to compute the sensitivity and specificity. At the threshold corresponding to peak F1 score, Resnet 18 had the highest sensitivity (0.638 ± 0.048) and specificity (0.950 ± 0.013), while VGG had the lowest (0.576 ± 0.040 sensitivity, 0.941 ± 0.012 specificity). These results are summarized in Table 3 and the average rate of false negative, false postivie, true negative, and true positive for each architecture are summarized in Table 4.

**Table 3:** AUC, optimal F1 score, sensitivity, and specificity for each network architecture in the testing set. Metrics are reported as a mean over the five trained networks per architecture ± standard deviation. Sensitivity and specificity are calculated at the threshold corresponding to the peak F1 score.

| Architecture | AUC | F1 Score | Sensitivity | Specificity |
| --- | --- | --- | --- | --- |
| Resnet 18 | $0.917 \pm 0.001$ | $0.586 \pm 0.010$ | $0.638 \pm 0.048$ | $0.950 \pm 0.013$ |
| Resnet 50 | $0.913 \pm 0.004$ | $0.560 \pm 0.013$ | $0.624 \pm 0.024$ | $0.944 \pm 0.006$ |
| Alexnet | $0.909 \pm 0.003$ | $0.569 \pm 0.021$ | $0.614 \pm 0.060$ | $0.949 \pm 0.019$ |
| densenet 121 | $0.907 \pm 0.003$ | $0.535 \pm 0.016$ | $0.597 \pm 0.038$ | $0.942 \pm 0.004$ |
| squeezenet | $0.904 \pm 0.005$ | $0.546 \pm 0.019$ | $0.602 \pm 0.042$ | $0.944 \pm 0.010$ |
| vgg 11 | $0.902 \pm 0.004$ | $0.520 \pm 0.010$ | $0.576 \pm 0.040$ | $0.941 \pm 0.012$ |

| Variable | Test N=2486 | Train N=22382 | P value | x Missing |
| --- | --- | --- | --- | --- |
| **Coronary Artery Disease** | 816 (34.52%) | 7250 (34.06%) | 0.65 | 1216 |
| **Myocardial Infarction** | 531 (22.46%) | 4672 (21.95%) | 0.57 | 1216 |
| **Congestive Heart Failure** | 512 (21.66%) | 4555 (21.4%) | 0.77 | 1216 |
| **Cerebrovascular Disease** | 463 (19.59%) | 4183 (19.65%) | 0.94 | 1216 |
| **Stroke or Transient Ischemic Attack** | 364 (15.4%) | 3239 (15.22%) | 0.81 | 1216 |
| **Atrial Fibrillation** | 318 (12.79%) | 3031 (13.54%) | 0.30 | 0 |
| **Body Mass Index:** | 30.00 (7.64) | 29.85 (7.75) | 0.41 | 4379 |
| **LVEF VALUE** | 57.70 (13.96) | 57.65 (14.05) | 0.93 | 18136 |

**Table 4:** Counts for false positive, false negative, true positive, and true negative for each network architecture.

| Class | False Negative | False Positive | True Negative | True Positive |
| --- | --- | --- | --- | --- |
| Resnet 18 | 68 (2.7%) | 129 (5.1%) | 2147 (86.4%) | 142 (5.7%) |
| Resnet50 | 81 (3.3%) | 112 (4.5%) | 2164 (87.0%) | 129 (5.2%) |
| Alexnet | 90 (3.6%) | 88 (3.5%) | 2188 (88.0%) | 120 (4.8%) |
| densenet121 | 90 (3.6%) | 131 (5.3%) | 2145 (86.3%) | 120 (4.8%) |
| squeezenet1 0 | 93 (3.7%) | 116 (4.7%) | 2160 (86.9%) | 117 (4.7%) |
| vgg11 | 90 (3.6%) | 126 (5.1%) | 2150 (86.5%) | 120 (4.8%) |

**Figure 2:**
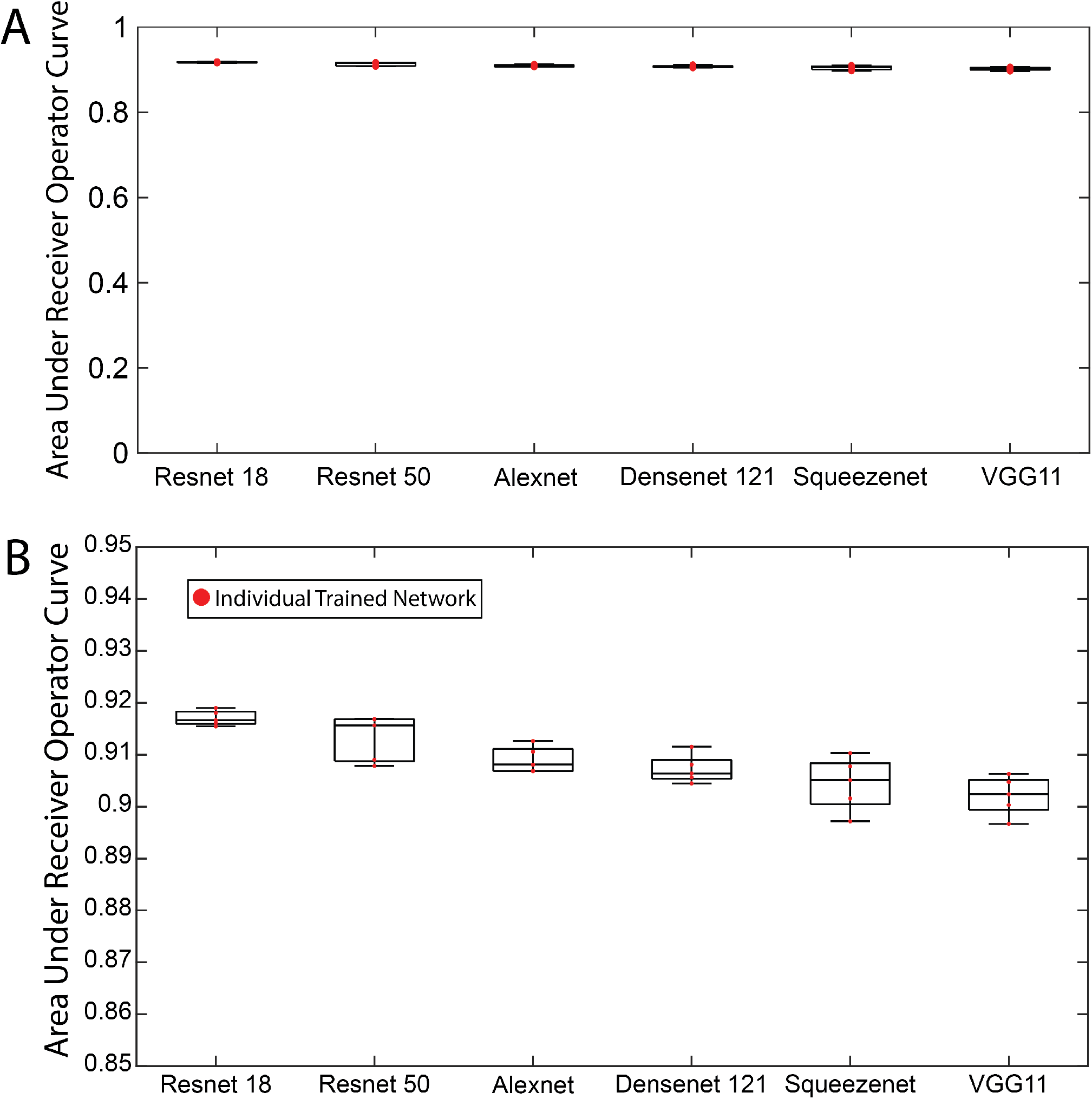
Area Under the Curve (AUC) metrics for each network tested. AUCs were calculated for the five training tests performed with different testing and training data. Panel B shows a zoomed in Y scale version of panel A.

### 4.2 Effects of Baseline Patient Characteristics

For each AI-ML architecture (Resnet 18, Resnet 50, Alexnet, Densenet 121, Squeezenet, and VGG), the highest-performing instance of the five trained instances was selected and thresholded based on the maximum F1 criterion as described above. Corresponding demographic and comorbidity data were compared for each AI-ML architecture between the correct and incorrect LVEF classifications. Table 5 shows the patient characteristics in the correct vs. incorrect for the best performing Resnet 18 architecture. The comparisons for each of the other networks can be found in the supplemental material (Tables S.1, through S.5). Statistical significance indicates a difference between comorbidity or demographic frequencies in correct vs. incorrect groups.

**Table 5:**
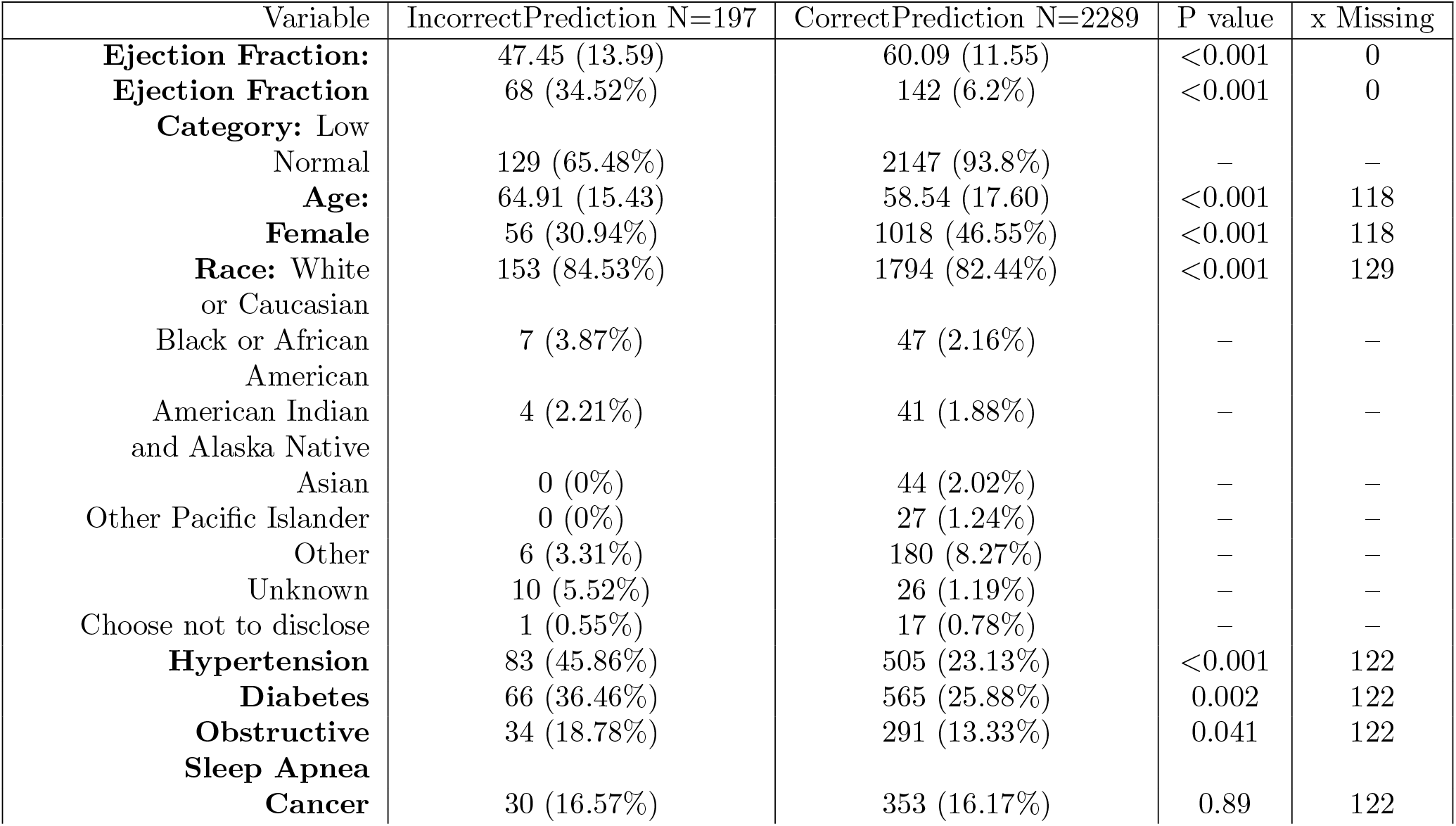
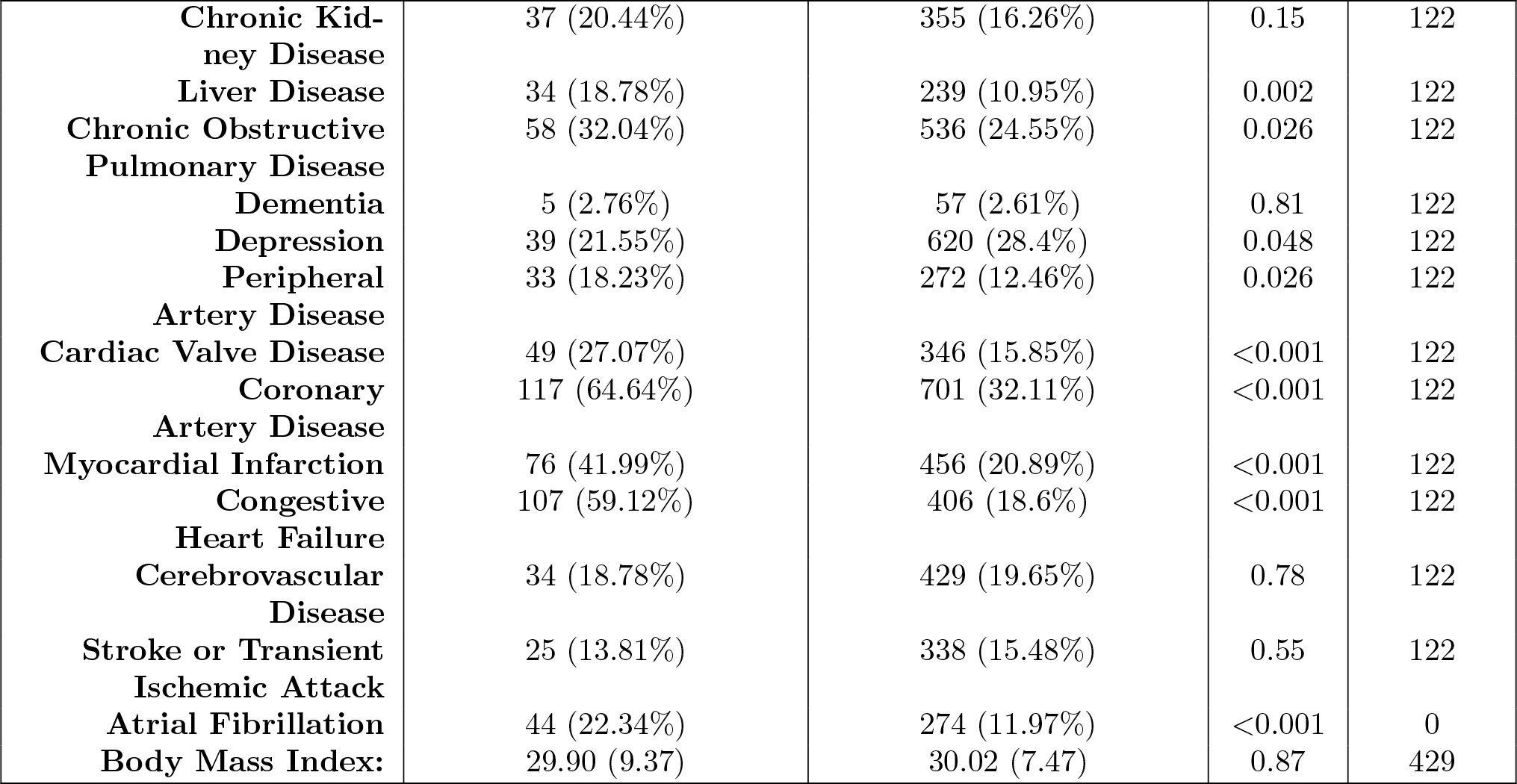
Demographic and Comorbidity comparison between correct predictions (true positive or true negative) vs incorrect predictions (false positive or false negative) for the best performing Resnet 18 implementation. Statistical significance indicates a difference between correct and incorrect.

The P-values for variable comparisons between correct and incorrect LVEF classification groups are shown as a heat-map in Table 6, where blue indicates a statistically significantly larger value (percentage for binary variables or mean for scalar variables) in the correct prediction group. Red indicates a statistically significantly larger value in the incorrect prediction group.

**Table 6:**
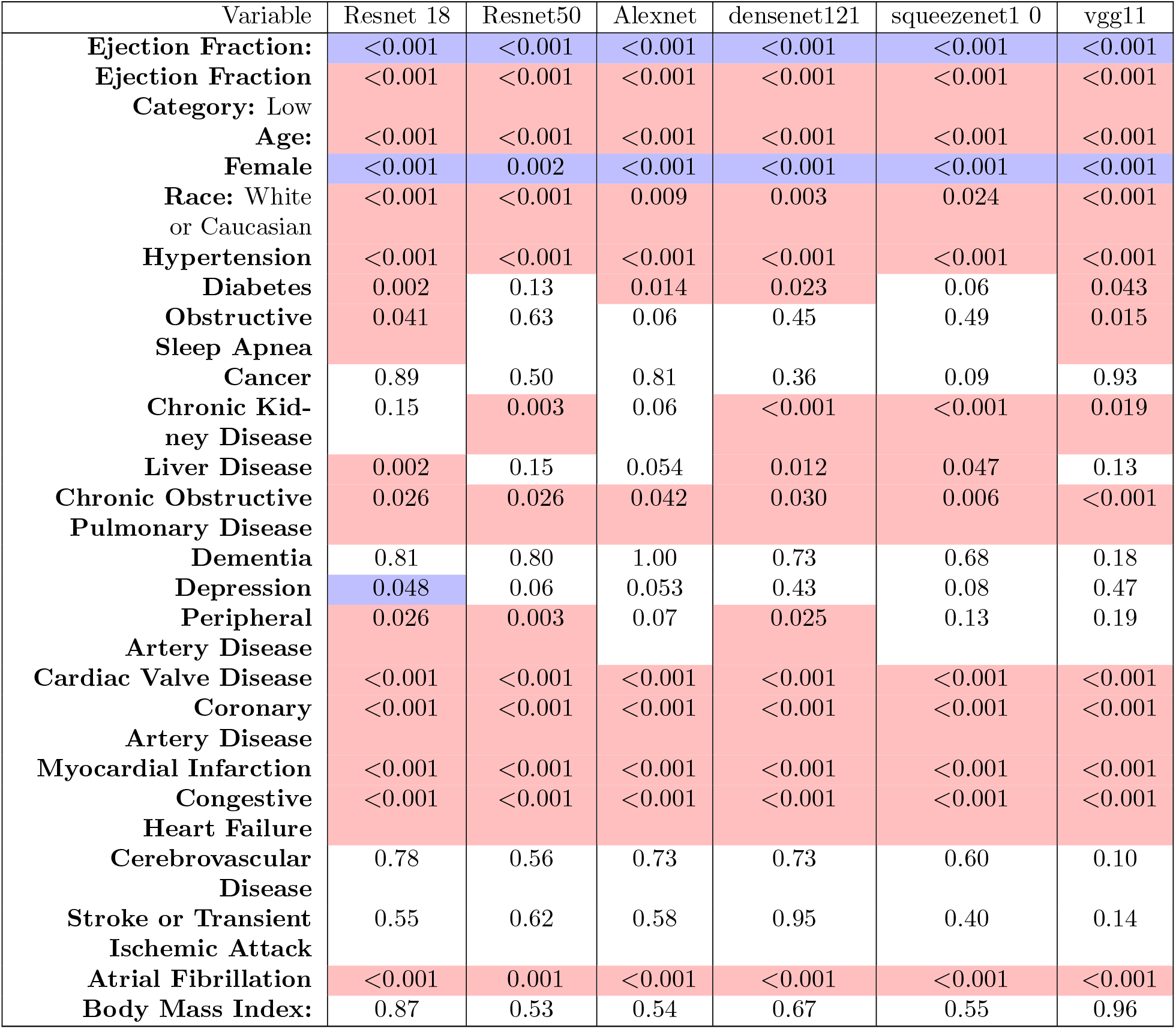
P values for each variuable across each architecture comparing correct predicitions (true positive or true negative) vs incorrect predictions (false positive or false negative). When significant (p ¡ 0.05), red indicates either higher percent or higher mean in the incorrect predictions, while blue indicates higher percent or hgigher mean in the correct predictions

## 5 Discussion and Conclusions

In summary, we report the first implementation and training of six “off-the-shelf” open-source AI-ML algorithms for use on ECG data to predict low or normal LVEF. There are three major findings from these implementations: (1) Implementing these architectures was relatively simple; (2) these architectures performed favorably compared with custom-built task-specific algorithms; and (3) despite excellent overall performance, we found that some patient characteristics were more associated with AI-ML LVEF misclassification and may have implications for downstream bias.

We showed that these open-source AI-ML architectures could be rapidly adapted and trained on realworld ECG data to perform a clinically valuable task previously demonstrated primarily by /emphcustom-built networks. We implemented AI-ML architectures using open-source packages available in the Python computing language, with minimal overhead and data manipulation. Furthermore, the computational resources necessary to train and test these networks were modest (single graphics card with 24 GB video memory, 20 CPU processor cores, 256 GB RAM). Additionally, minimal data manipulation and preprocessing were required to fit the AI-ML architecture prespecified input and output parameters. The application of AI-ML technology to ECG data may provide important diagnostic value to low healthcare resource environments – the use of widely-available, open-source architectures that do not require substantial computing power is an important component for such a deployment.

We also showed that open-source AI-ML approaches are consistent with the performance of many of the custom-built approaches on an identical task when comparing routine AI-ML metrics. Resnet 18 performed the best with a mean AUC of 0.917 ± 0.001 over five instances, with VGG11 performing least accurately but still highly successful with an average AUC of 0.902 ± 0.004. These values are comparable to other published architectures [4], [6], [7]. Furthermore, the max F1-score was also relatively high across networks, with the mean F1-score ranging from 0.586 to 0.520 for the Resnet 18 and VGG 11 networks, respectively. Interestingly, the sensitivity ranged from 58% to 63% and specificity from 94% to 95% at the maximum F1-score threshold per network. The clinical implications of the relatively low sensitivity and high specificity indicate that our current implementation is not an ideal “screening” test but could be used to rule in low LVEF. Adjustments to the selected threshold would allow for tuning of the network towards higher sensitivity or specificity.

The metrics of AUC, F1-score, sensitivity, and specificity are commonly used to evaluate AI-ML algorithms, particularly in non-clinical settings. However, in our results there is little absolute variability in these measures among the tested architectures: less than 0.03 in AUC, 0.07 in F1-score, 5% in sensitivity, and 1% in specificity. Therefore, selecting an ideal *clinical* AI-ML approach based on conventional metrics alone may not be helpful – at least, when they are each so close. Other criteria, such as resource utilization and portability may become important. Furthermore, these metrics convey minimal clinically relevant information. Measures to date do not provide information about how the AI-ML approaches perform with different cohorts of patients and if particular patient characteristics are associated with worse AI-ML performance – potential contributors to bias and worsening disparities of care. To provide a framework to understand these possibilities, we investigated whether baseline characteristics differed in patients with correct vs. incorrect AI-ML LVEF classification.

We found several baseline comorbidities that were associated with were statistically different in patients with correct vs. incorrect AI-ML LVEF classification. These include, hypertension (HTN), chronic obstructive pulmonary disease (COPD), coronary artery disease (CAD), myocardial infarction (MI), valve disease, congestive heart failure (CHF), and atrial fibrillation (AF). Physiologic reasons for this poor performance can be postulated for patients with an intrinsic cardiac pathology such as HTN, CHF, MI, CAD, valvular disease, or AF because each can significantly affect the cardiac electrical conduction system and alter the 12-lead ECG. However, COPD is not an intrinsic cardiac pathology but could be related to changes in cardiac electrical signals. Patients with COPD often have higher lung volumes which can be an excellent electrical insulator. These hypothesis are preliminary and not explicitly confirmed by the data presented, but simply demonstrate the possible links between changes in ECG signals and baseline patient comorbidities.

Importantly, we also found constitutional patient characteristics, such as gender and race, where performance differed. Specifically, patients who are older and White are more likely to have a false positive or false negative AI-ML classified low-LVEF. We realize that these results are not definitive performance metrics, but the demonstrate the massive potential for AI-ML approaches to harbor bias and impact disparities of care positively or negatively. AI-ML approaches used in other fields have begun to grapple with these realities of bias. We find it prudent to highlight that AI-ML applied to ECG related problems could also suffer from similar inherent biases. Further work is needed to capture, describe, and correct for the inevitable disparities that will develop from these approaches.

Additionally, our results will contribute to explanatory AI-ML. Traditional, human-based ECG interpretation has been refined over decades, to describe patterns associated with disease commonly via pathophysiologic links. In contrast, modern ML-ECG algorithms remain more ‘black-box’ technologies that generate predictive output with little explanation as to why the algorithm has linked a specific ECG to a target. And while some algorithms intuitively link the ECG to a related cardiovascular outcome (e.g., future arrhythmia),[13], [21] others have linked the ECG waveform to seemingly-unrelated conditions such as liver disease – a less clear pathophysiologic link [22]. Future studies could use explanatory machine learning techniques to elucidate what areas of the ECG are identified by each architecture and if changes in those regions of the ECG are related to an underlying diagnosis [23]–[25].

One crucial factor that could be driving these results is the frequency of patients with specific underlying characteristics or comorbidities in the training data. However, our patient population has average or higher rates of comorbidities than the general US population [26]–[28]. Our data also have higher rates of cardiac diagnoses, including CAD, MI, AF, and others. Furthermore, our training vs. testing dataset had minimal differences in patient baseline characteristics and comorbidities.

There were several limitations to this study. First, these ML architectures we implemented were designed for use with images, and thus in some cases we were required to reshape our input ECGs to allow for application of these architectures (Densenet 121 and VGG11). Such reshaping may be deleterious for the spatially coherent information present in an ECG signal, and thus may negatively impact network performance. Furthermore, such open-source AI-ML architectures are not designed to leverage the unique features of ECG data such as its temporal coherence, as is seen in other ECG specific approaches [1], [2]. Our dataset was also limited to a single center with a relatively socially homogenous population. Finally, our dataset was biased to have more individuals with a normal LVEF.

In conclusion, we found that several “off-the-shelf,” open-source AI-ML architectures could be used to predict low LVEF from ECGs. Specifically, we found these approaches were easy to implement and performed comparably to previously reported custom-built networks. Furthermore, we found baseline patient characteristics differed substantially between patients with correct versus incorrect AI-ML LVEF classification. These findings should be considered in the pursuit of efficient and equitable deployment of AI-ML technologies moving forward.

## Data Availability

Data from this study is not publicly available. Data may be made available via collaboration agreement upon reasonable and approved request to the authors.

## 5.1 Acknowledgments

Support for this research came from the Center for Integrative Biomedical Computing (www.sci.utah.edu/cibc), NIH/NIGMS grants P41 GM103545 and R24 GM136986, NIH/NIBIB grant U24EB029012, National Heart, Lung, And Blood Institute of the National Institutes of Health under Award Number K23HL143156 (to BAS) and Award Number F30HL149327 (to BZ) and the Nora Eccles Harrison Foundation for Cardiovascular Research.

## 6 Supplemental

The following data is present as supplemental to the primary manuscript. This supplemental data consists of demographic and co-morbidity tables for the best trained instance for each architecture. Additionally training time is reported for each network architecture.

**Table S.1:** Demographic and Comorbidity comparison between correct predicitions (true positive or true negative) vs incorrect predictions (false positive or false negative) for the best performing Resnet50 implementation. Statistical significance indicates a difference between correct and incorrect.

| Variable | IncorrectPrediction N=197 | CorrectPrediction N=2289 | P value | x Missing |
| --- | --- | --- | --- | --- |
| **Ejection Fraction:** | 46.24 (15.00) | 60.17 (11.30) | <0.001 | 0 |
| **Ejection Fraction** | 81 (41.97%) | 129 (5.63%) | <0.001 | 0 |
| **Category:** Low |  |  |  |  |
| Normal | 112 (58.03%) | 2164 (94.37%) | – | – |
| **Age:** | 65.40 (15.62) | 58.52 (17.57) | <0.001 | 118 |
| **Female** | 59 (33.91%) | 1015 (46.26%) | 0.002 | 118 |
| **Race:** White | 151 (86.78%) | 1796 (82.27%) | <0.001 | 129 |
| or Caucasian |  |  |  |  |
| Black or African | 4 (2.3%) | 50 (2.29%) | – | – |
| American |  |  |  |  |
| American Indian | 3 (1.72%) | 42 (1.92%) | – | – |
| and Alaska Native |  |  |  |  |
| Asian | 0 (0%) | 44 (2.02%) | – | – |
| Other Pacific Islander | 0 (0%) | 27 (1.24%) | – | – |
| Other | 6 (3.45%) | 180 (8.25%) | – | – |
| Unknown | 10 (5.75%) | 26 (1.19%) | – | – |
| Choose not to disclose | 0 (0%) | 18 (0.82%) | – | – |
| **Hypertension** | 77 (44.25%) | 511 (23.33%) | <0.001 | 122 |
| **Diabetes** | 55 (31.61%) | 576 (26.3%) | 0.13 | 122 |
| **Obstructive** | 26 (14.94%) | 299 (13.65%) | 0.63 | 122 |
| **Sleep Apnea** |  |  |  |  |
| **Cancer** | 25 (14.37%) | 358 (16.35%) | 0.50 | 122 |
| **Chronic Kid-** | 43 (24.71%) | 349 (15.94%) | 0.003 | 122 |
| **ney Disease** |  |  |  |  |
| **Liver Disease** | 26 (14.94%) | 247 (11.28%) | 0.15 | 122 |
| **Chronic Obstructive** | 56 (32.18%) | 538 (24.57%) | 0.026 | 122 |
| **Pulmonary Disease** |  |  |  |  |
| **Dementia** | 5 (2.87%) | 57 (2.6%) | 0.80 | 122 |
| **Depression** | 38 (21.84%) | 621 (28.36%) | 0.06 | 122 |
| **Peripheral** | 35 (20.11%) | 270 (12.33%) | 0.003 | 122 |
| **Artery Disease** |  |  |  |  |
| **Cardiac Valve Disease** | 49 (28.16%) | 346 (15.8%) | <0.001 | 122 |
| **Coronary** | 109 (62.64%) | 709 (32.37%) | <0.001 | 122 |
| **Artery Disease** |  |  |  |  |
| **Myocardial Infarction** | 67 (38.51%) | 465 (21.23%) | <0.001 | 122 |
| **Congestive** | 99 (56.9%) | 414 (18.9%) | <0.001 | 122 |
| **Heart Failure** |  |  |  |  |
| **Cerebrovascular** | 37 (21.26%) | 426 (19.45%) | 0.56 | 122 |
| **Disease** |  |  |  |  |
| **Stroke or Transient Ischemic Attack** | 29 (16.67%) | 334 (15.25%) | 0.62 | 122 |
| **Atrial Fibrillation** | 39 (20.21%) | 279 (12.17%) | 0.001 | 0 |
| **Body Mass Index:** | 29.53 (10.25) | 30.05 (7.39) | 0.53 | 429 |

**Table S.2:** Demographic and Comorbidity comparison between correct predicitions (true positive or true negative) vs incorrect predictions (false positive or false negative) for the best performing Alexnet implementation. Statistical significance indicates a difference between correct and incorrect.

| Variable | IncorrectPrediction N=197 | CorrectPrediction N=2289 | P value | x Missing |
| --- | --- | --- | --- | --- |
| **Ejection Fraction:** | 43.56 (14.30) | 60.29 (11.17) | <0.001 | 0 |
| **Ejection Fraction Category: Low** | 90 (50.56%) | 120 (5.2%) | <0.001 | 0 |
| Normal | 88 (49.44%) | 2188 (94.8%) | – | – |
| **Age:** | 65.60 (14.64) | 58.55 (17.62) | <0.001 | 118 |
| **Female** | 47 (29.38%) | 1027 (46.51%) | <0.001 | 118 |
| **Race: White or Caucasian** | 136 (85%) | 1811 (82.43%) | 0.009 | 129 |
| Black or African American | 5 (3.12%) | 49 (2.23%) | – | – |
| American Indian and Alaska Native | 2 (1.25%) | 43 (1.96%) | – | – |
| Asian | 1 (0.62%) | 43 (1.96%) | – | – |
| Other Pacific Islander | 1 (0.62%) | 26 (1.18%) | – | – |
| Other | 7 (4.38%) | 179 (8.15%) | – | – |
| Unknown | 8 (5%) | 28 (1.27%) | – | – |
| Choose not to disclose | 0 (0%) | 18 (0.82%) | – | – |
| **Hypertension** | 75 (46.88%) | 513 (23.28%) | <0.001 | 122 |
| **Diabetes** | 56 (35%) | 575 (26.09%) | 0.014 | 122 |
| **Obstructive Sleep Apnea** | 30 (18.75%) | 295 (13.38%) | 0.06 | 122 |
| **Cancer** | 27 (16.88%) | 356 (16.15%) | 0.81 | 122 |
| **Chronic Kidney Disease** | 35 (21.88%) | 357 (16.2%) | 0.06 | 122 |
| **Liver Disease** | 26 (16.25%) | 247 (11.21%) | 0.054 | 122 |
| **Chronic Obstructive Pulmonary Disease** | 51 (31.87%) | 543 (24.64%) | 0.042 | 122 |
| **Dementia** | 4 (2.5%) | 58 (2.63%) | 1.00 | 122 |
| **Depression** | 34 (21.25%) | 625 (28.36%) | 0.053 | 122 |
| **Peripheral Artery Disease** | 28 (17.5%) | 277 (12.57%) | 0.07 | 122 |
| **Cardiac Valve Disease** | 55 (34.38%) | 340 (15.43%) | <0.001 | 122 |
| **Coronary Artery Disease** | 107 (66.88%) | 711 (32.26%) | <0.001 | 122 |
| **Myocardial Infarction** | 68 (42.5%) | 464 (21.05%) | <0.001 | 122 |
| **Congestive Heart Failure** | 101 (63.12%) | 412 (18.69%) | <0.001 | 122 |
| **Cerebrovascular Disease** | 33 (20.62%) | 430 (19.51%) | 0.73 | 122 |
| **Stroke or Transient Ischemic Attack** | 27 (16.88%) | 336 (15.25%) | 0.58 | 122 |
| **Atrial Fibrillation** | 41 (23.03%) | 277 (12%) | <0.001 | 0 |
| **Body Mass Index:** | 29.54 (9.77) | 30.04 (7.46) | 0.54 | 429 |

**Table S.3:** Demographic and Comorbidity comparison between correct predicitions (true positive or true negative) vs incorrect predictions (false positive or false negative) for the best performing squeezenet10 implementation. Statistical significance indicates a difference between correct and incorrect.

| Variable | IncorrectPrediction N=197 | CorrectPrediction N=2289 | P value | x Missing |
| --- | --- | --- | --- | --- |
| **Ejection Fraction:** | 45.47 (15.05) | 60.34 (11.11) | <0.001 | 0 |
| **Ejection Fraction Category: Low** | 93 (44.5%) | 117 (5.14%) | <0.001 | 0 |
| Normal | 116 (55.5%) | 2160 (94.86%) | – | – |
| **Age:** | 64.35 (16.25) | 58.55 (17.55) | <0.001 | 118 |
| **Female** | 63 (32.31%) | 1011 (46.53%) | <0.001 | 118 |
| **Race: White or Caucasian** | 164 (84.54%) | 1783 (82.43%) | 0.024 | 129 |
| Black or African American | 6 (3.09%) | 48 (2.22%) | – | – |
| American Indian and Alaska Native | 4 (2.06%) | 41 (1.9%) | – | – |
| Asian | 1 (0.52%) | 43 (1.99%) | – | – |
| Other Pacific Islander | 0 (0%) | 27 (1.25%) | – | – |
| Other | 10 (5.15%) | 176 (8.14%) | – | – |
| Unknown | 8 (4.12%) | 28 (1.29%) | – | – |
| Choose not to disclose | 1 (0.52%) | 17 (0.79%) | – | – |
| **Hypertension** | 93 (47.69%) | 495 (22.82%) | <0.001 | 122 |
| **Diabetes** | 63 (32.31%) | 568 (26.19%) | 0.06 | 122 |
| **Obstructive Sleep Apnea** | 30 (15.38%) | 295 (13.6%) | 0.49 | 122 |
| **Cancer** | 40 (20.51%) | 343 (15.81%) | 0.09 | 122 |
| **Chronic Kidney Disease** | 55 (28.21%) | 337 (15.54%) | <0.001 | 122 |
| **Liver Disease** | 31 (15.9%) | 242 (11.16%) | 0.047 | 122 |
| **Chronic Obstructive Pulmonary Disease** | 65 (33.33%) | 529 (24.39%) | 0.006 | 122 |
| **Dementia** | 6 (3.08%) | 56 (2.58%) | 0.68 | 122 |
| **Depression** | 44 (22.56%) | 615 (28.35%) | 0.08 | 122 |
| **Peripheral Artery Disease** | 32 (16.41%) | 273 (12.59%) | 0.13 | 122 |
| **Cardiac Valve Disease** | 60 (30.77%) | 335 (15.44%) | <0.001 | 122 |
| **Coronary Artery Disease** | 122 (62.56%) | 696 (32.09%) | <0.001 | 122 |
| **Myocardial Infarction** | 84 (43.08%) | 448 (20.65%) | <0.001 | 122 |
| **Congestive Heart Failure** | 114 (58.46%) | 399 (18.4%) | <0.001 | 122 |
| **Cerebrovascular Disease** | 41 (21.03%) | 422 (19.46%) | 0.60 | 122 |
| **Stroke or Transient Ischemic Attack** | 34 (17.44%) | 329 (15.17%) | 0.40 | 122 |
| **Atrial Fibrillation** | 52 (24.88%) | 266 (11.68%) | <0.001 | 0 |
| **Body Mass Index:** | 29.62 (9.31) | 30.05 (7.47) | 0.55 | 429 |

**Table S.4:** Demographic and Comorbidity comparison between correct predicitions (true positive or true negative) vs incorrect predictions (false positive or false negative) for the best performing densenet121 implementation. Statistical significance indicates a difference between correct and incorrect.

| Variable | IncorrectPrediction N=197 | CorrectPrediction N=2289 | P value | x Missing |
| --- | --- | --- | --- | --- |
| **Ejection Fraction:** | 46.41 (15.46) | 60.33 (11.10) | <0.001 | 0 |
| **Ejection Fraction Category: Low** | 90 (40.72%) | 120 (5.3%) | <0.001 | 0 |
| Normal | 131 (59.28%) | 2145 (94.7%) | – | – |
| **Age:** | 65.37 (16.13) | 58.44 (17.53) | <0.001 | 118 |
| **Female** | 65 (32.5%) | 1009 (46.54%) | <0.001 | 118 |
| **Race: White or Caucasian** | 169 (84.92%) | 1778 (82.39%) | 0.003 | 129 |
| Black or African American | 7 (3.52%) | 47 (2.18%) | – | – |
| American Indian and Alaska Native | 3 (1.51%) | 42 (1.95%) | – | – |
| Asian | 0 (0%) | 44 (2.04%) | – | – |
| Other Pacific Islander | 0 (0%) | 27 (1.25%) | – | – |
| Other | 9 (4.52%) | 177 (8.2%) | – | – |
| Unknown | 9 (4.52%) | 27 (1.25%) | – | – |
| Choose not to disclose | 2 (1.01%) | 16 (0.74%) | – | – |
| **Hypertension** | 93 (46.5%) | 495 (22.87%) | <0.001 | 122 |
| **Diabetes** | 67 (33.5%) | 564 (26.06%) | 0.023 | 122 |
| **Obstructive Sleep Apnea** | 31 (15.5%) | 294 (13.59%) | 0.45 | 122 |
| **Cancer** | 37 (18.5%) | 346 (15.99%) | 0.36 | 122 |
| **Chronic Kidney Disease** | 52 (26%) | 340 (15.71%) | <0.001 | 122 |
| **Liver Disease** | 34 (17%) | 239 (11.04%) | 0.012 | 122 |
| **Chronic Obstructive Pulmonary Disease** | 63 (31.5%) | 531 (24.54%) | 0.030 | 122 |
| **Dementia** | 6 (3%) | 56 (2.59%) | 0.73 | 122 |
| **Depression** | 51 (25.5%) | 608 (28.1%) | 0.43 | 122 |
| **Peripheral Artery Disease** | 36 (18%) | 269 (12.43%) | 0.025 | 122 |
| **Cardiac Valve Disease** | 65 (32.5%) | 330 (15.25%) | <0.001 | 122 |
| **Coronary Artery Disease** | 129 (64.5%) | 689 (31.84%) | <0.001 | 122 |
| **Myocardial Infarction** | 79 (39.5%) | 453 (20.93%) | <0.001 | 122 |
| **Congestive Heart Failure** | 116 (58%) | 397 (18.35%) | <0.001 | 122 |
| **Cerebrovascular Disease** | 41 (20.5%) | 422 (19.5%) | 0.73 | 122 |
| **Stroke or Transient Ischemic Attack** | 31 (15.5%) | 332 (15.34%) | 0.95 | 122 |
| **Atrial Fibrillation** | 57 (25.79%) | 261 (11.52%) | <0.001 | 0 |
| **Body Mass Index:** | 30.29 (9.54) | 29.98 (7.43) | 0.67 | 429 |

**Table S.5:** Demographic and Comorbidity comparison between correct predicitions (true positive or true negative) vs incorrect predictions (false positive or false negative) for the best performing vgg11 implementation. Statistical significance indicates a difference between correct and incorrect.

| Variable | IncorrectPrediction N=197 | CorrectPrediction N=2289 | P value | x Missing |
| --- | --- | --- | --- | --- |
| **Ejection Fraction:** | 45.62 (14.65) | 60.37 (11.14) | <0.001 | 0 |
| **Ejection Fraction Category: Low** | 90 (41.67%) | 120 (5.29%) | <0.001 | 0 |
| Normal | 126 (58.33%) | 2150 (94.71%) | – | – |
| **Age:** | 64.20 (16.13) | 58.56 (17.57) | <0.001 | 118 |
| **Female** | 66 (33.67%) | 1008 (46.41%) | <0.001 | 118 |
| **Race: White or Caucasian** | 171 (87.24%) | 1776 (82.18%) | <0.001 | 129 |
| Black or African American | 4 (2.04%) | 50 (2.31%) | – | – |
| American Indian and Alaska Native | 3 (1.53%) | 42 (1.94%) | – | – |
| Asian | 0 (0%) | 44 (2.04%) | – | – |
| Other Pacific Islander | 1 (0.51%) | 26 (1.2%) | – | – |
| Other | 6 (3.06%) | 180 (8.33%) | – | – |
| Unknown | 10 (5.1%) | 26 (1.2%) | – | – |
| Choose not to disclose | 1 (0.51%) | 17 (0.79%) | – | – |
| **Hypertension** | 91 (46.67%) | 497 (22.91%) | <0.001 | 122 |
| **Diabetes** | 64 (32.82%) | 567 (26.14%) | 0.043 | 122 |
| **Obstructive Sleep Apnea** | 38 (19.49%) | 287 (13.23%) | 0.015 | 122 |
| **Cancer** | 32 (16.41%) | 351 (16.18%) | 0.93 | 122 |
| **Chronic Kidney Disease** | 44 (22.56%) | 348 (16.04%) | 0.019 | 122 |
| **Liver Disease** | 29 (14.87%) | 244 (11.25%) | 0.13 | 122 |
| **Chronic Obstructive Pulmonary Disease** | 69 (35.38%) | 525 (24.2%) | <0.001 | 122 |
| **Dementia** | 8 (4.1%) | 54 (2.49%) | 0.18 | 122 |
| **Depression** | 50 (25.64%) | 609 (28.08%) | 0.47 | 122 |
| **Peripheral Artery Disease** | 31 (15.9%) | 274 (12.63%) | 0.19 | 122 |
| **Cardiac Valve Disease** | 57 (29.23%) | 338 (15.58%) | <0.001 | 122 |
| **Coronary Artery Disease** | 117 (60%) | 701 (32.32%) | <0.001 | 122 |
| **Myocardial Infarction** | 76 (38.97%) | 456 (21.02%) | <0.001 | 122 |
| **Congestive Heart Failure** | 116 (59.49%) | 397 (18.3%) | <0.001 | 122 |
| **Cerebrovascular Disease** | 47 (24.1%) | 416 (19.18%) | 0.10 | 122 |
| **Stroke or Transient Ischemic Attack** | 37 (18.97%) | 326 (15.03%) | 0.14 | 122 |
| **Atrial Fibrillation** | 44 (20.37%) | 274 (12.07%) | <0.001 | 0 |
| **Body Mass Index:** | 29.97 (9.52) | 30.01 (7.45) | 0.96 | 429 |

**Table S.6:**
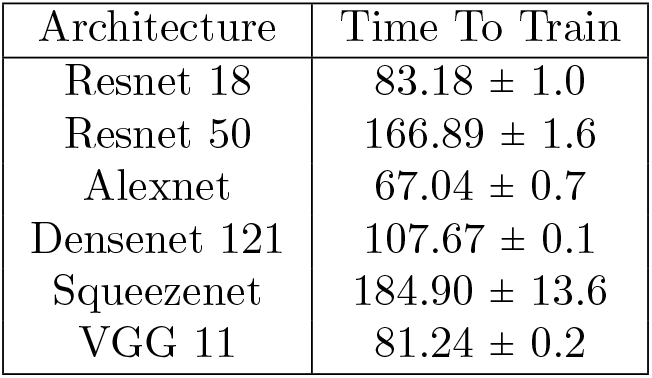
Average time to complete the full training regimen in minutes. Times are reported as an average plus or minus one standard deviation over the five instances per network architecture.

| Architecture | Time To Train |
| --- | --- |
| Resnet 18 | $83.18 \pm 1.0$ |
| Resnet 50 | $166.89 \pm 1.6$ |
| Alexnet | $67.04 \pm 0.7$ |
| Densenet 121 | $107.67 \pm 0.1$ |
| Squeezenet | $184.90 \pm 13.6$ |
| VGG 11 | $81.24 \pm 0.2$ |

